# Linking brain networks to cognition with magnetoencephalography in paediatric autoimmune encephalitis

**DOI:** 10.1101/2024.04.04.24305194

**Authors:** Charly Billaud, Amanda G. Wood, Daniel Griffiths-King, Klaus Kessler, Evangeline Wassmer, Elaine Foley, Sukhvir K. Wright

## Abstract

Paediatric autoimmune encephalitis (e.g., acute disseminated encephalomyelitis, N-methyl-D-aspartate receptor antibody encephalitis) is an inflammatory brain disease that causes cognitive deficits, psychiatric symptoms, seizures, MRI, and EEG abnormalities. Patients can continue to experience residual cognitive difficulties months to years after the acute illness. Magnetoencephalography (MEG) can examine neural changes in the absence of frank structural abnormalities and may help identify factors predicting children at risk of long-term cognitive deficits. We predicted that theta and delta brain functional connectivity networks would be associated with processing speed and working memory in children with autoimmune encephalitis.

Participants were children diagnosed with autoimmune encephalitis at least 18 months before testing and typically developing children. All completed MEG recording (Elekta Neuromag Triux) at rest, eyes open with a fixation cross during six minutes; T1 MRI scans; and cognitive evaluation using the primary subtests of the Weschler Intelligence Scale for Children, fifth edition. Brain connectivity, specifically in delta and theta brain activity, was estimated with amplitude envelope correlation, and network efficiency was measured using graph measures (global efficiency, local efficiency, modularity). The measures were compared across the two groups with permutation correction for multiple thresholds. Finally, statistical associations with processing speed and working memory scores were tested in the autoimmune encephalitis group.

Age and sex-matched cohorts of 12 children with AE (11.2±3.5y, IQR 9y; 5M:7F) and 12 typically developing controls (10.6±3.2y, IQR 7y; 8M:4F) participated in this study. On average, children with autoimmune encephalitis did not differ from controls in working memory (*t*(21)= 1.449; *p* = .162; *d* = 0.605) but had a significantly lower processing speed (*t*(21) = 2.463; *p* = .023; Cohen’s *d* = 1.028). The groups did not differ in theta network topology measures but the autoimmune encephalitis group had a significantly lower delta local efficiency across all thresholds tested (*d* = −1.60 at network threshold 14%). Theta modularity was associated with lower working memory (*β* = -.781; *t*(8) = −2.588, *p* = .032) but this effect did not survive correction for multiple comparisons (*p(corr)* = .224). No other graph measure was significantly associated with psychometric scores in the autoimmune encephalitis group.

MEG was able to capture network alterations in paediatric autoimmune encephalitis patients, specifically in the topological organisation of delta brain activity. This preliminary study demonstrates that MEG is an appropriate tool for assessing children with autoimmune encephalitis; future studies should focus on confirming which functional networks can predict cognitive performance.

## Introduction

Paediatric auto-immune encephalitis (PAE) is an inflammatory brain disease associated with seizures, movement disorders, neuropsychiatric symptoms, and cognitive deficits^1–4^. Despite advances in diagnosing and treating PAE, a proportion of children are still left with long-term cognitive and academic difficulties (up to 45% in N-methyl-D-aspartate receptor antibody encephalitis (NMDARE)^5–7^ and 15.8–22% in acute disseminated encephalomyelitis (ADEM)^8, 9^). Routinely used clinical outcome assessments such as the modified Rankin Scale (mRS) or the Extended Disability Status Scale (EDSS) focus on motor abilities missing independent subtle cognitive deficits^6, 10–12^. Identifying factors that predict neuropsychological outcome would allow rehabilitation therapies to be focussed on PAE patients at highest risk of cognitive difficulties^5^. A recent review in adults with AE found the most frequent impairments and highest yield neuropsychological measures were found in tests of visual and verbal learning/memory, processing speed, attention, and executive functions.^13^

Functional connectivity is an indicator of neural activity that reveals a systematic relationship between brain areas and is therefore interpreted as reflecting neural connections. Resting state functional connectivity analyses that focus on the meaningful organisation of spontaneous brain activity, in the absence of any specific task (‘at-rest’), offer great potential for investigating the underlying mechanisms of cognitive sequelae in autoimmune encephalitis (AE). In adults, functional Magnetic Resonance Imaging (fMRI) studies suggest that patients with AE have reduced functional connectivity at rest.^14^ Altered connectivity (i.e. “dysconnectivity”) may reflect disruptions in neural connections, leading to impairment of cognitive functions that rely on inter-area neural communication. Network dysconnectivity in adult AE has been linked to lower cognitive performance^15–20^ disease severity^15, 16, 21^, higher mood lability^22^, and psychiatric symptoms^16^. Specifically, deficits in working memory and information processing speed significantly correlated with functional connectivity measures.^23^ One study in adult limbic encephalitis (a form of AE) hypothesised that damaged brain structures could provoke increased connectivity associated with deficits, reporting a positive relationship between functional connectivity to the insular cortex and impaired verbal episodic memory.^18^ To date, children and young people with AE have not been specifically investigated for functional dysconnectivity, despite evidence to show they may be more adversely affected in terms of neuropsychological and cognitive outcomes^5, 24^, in part due to the vulnerability of the developing brain.^25^ It is therefore imperative that paediatric neuroimaging investigations into autoimmune encephalitis are conducted in order to understand changes in functional connectivity that could predict negative developmental outcomes.

Magnetoencephalography (MEG) is an alternative non-invasive method used for measuring functional brain dynamics^26^, but studies in AE are limited to case reports.^27, 28^ MEG provides a direct measure of neuronal activity, unlike fMRI which relies on a surrogate signal based on blood oxygenation levels. MEG can reveal widespread changes in functional connectivity with higher temporal resolution than fMRI, which is especially relevant to examining synchrony of neural oscillations across brain regions^29^. In addition, MEG provides a better spatial resolution than EEG, being less susceptible to the spatial distortions in recorded brain activity caused by interactions between the signal and tissue layers with variable electric conductivity surrounding the brain^30^. Resting-state MEG recordings can be used to infer functional networks^31, 32^, in a similar way as fMRI in AE^16^. MEG has been widely used in paediatric epilepsy to predict cognitive outcomes^33^ and has the same advantages as EEG, i.e. it is cost-effective, non-invasive and reproducible^34^. In addition, MEG is practically preferrable for children, requiring less preparation time than EEG,^35^ and does not require children to lie still for long periods in a noisy environment like fMRI.

EEG is abnormal in >95% of adult and paediatric AE patients showing encephalopathy, but changes in specific frequency bands such as altered theta-delta activity, extreme delta brushes, and generalized rhythmic delta activity^36–39^ have also been widely reported.^36, 40^ Theta brain networks have been linked to attention and stimulus processing (sensation and perception), encoding and consolidation of information in memory, and executive functions including working memory^41^. Delta networks are also associated with attention and concentration and working memory^42^.

In this study, we investigated whether MEG-derived delta and theta connectivity in PAE was associated with cognitive measures in the long-term. We hypothesised that following AE, children would have lower scores in processing speed and working memory, as well as changes in delta and theta frequency resting-state network compared to typically developing controls. We further hypothesised that these network measures would be associated with lower cognitive performance in PAE.

## Materials and methods

### Participants

Patients were recruited between 2018 and 2022 from a single-centre, Birmingham Children’s Hospital, Birmingham UK as part of a larger study investigating the effect of neurological disease on the developing brain (Aston University Ethics reference #18/LO/0990; #IRAS 233424). Healthy, typically developing controls were recruited from the local community through social media advertisement and local outreach events (Aston University Ethics reference #HLS21011). Consent was obtained according to the Declaration of Helsinki and approved by ethical committees of each institution.

Patients were selected based upon the following criteria: Diagnosis of immune-mediated encephalitis, according to established criteria^1, 3^, at least 18 months after disease onset, aged 6 to 16 years at the time of recruitment to the study and cognitive assessment. Exclusion criteria included dissent of the child from participating and presence of contraindication for MRI scanning. For typically developing controls, exclusion criteria included a diagnosis of learning difficulty, psychiatric, neurodevelopmental, neurological disorder, a known or suspected cerebral abnormality, dissent of the child from participating and/or presence of contraindication for MRI scanning.

As an observational study where all participants followed the same protocol, participants were not randomized nor were researchers blinded to group membership.

Information regarding the children’s clinical and disease course were collected from medical records by paediatric neurologists (SW and EW) including clinical assessment of The Modified Rankin Scale (mRS) to classify the disability in patients. A score of 0 represents no disability, 3 is moderate disability requiring some help but able to walk and 5 is severe disability requiring constant care for all needs; 6 is death ^43^.

### Neuropsychological assessment

The *Wechsler Intelligence Scale for Children, 5th Edition* (WISC-V) was used to assess general intellectual functioning in all children from 6 years to 16 years and 11 months^44^. The Wechsler scale uses a battery of psychometric tests that assess intellectual quotient (IQ) into separate indices^44^. For the fifth edition of the WISC, the subtests are classified into the following indices: Verbal Comprehension, Visual Spatial, Fluid Reasoning, Working Memory, and Processing Speed^44^. Working Memory and Processing Speed composite scores were selected for this study analysis given the evidence that these are the most frequently studied and testing recommended in adult and paediatric AE and neuroinflammatory disease.^13, 45, 46^

### MRI image acquisition

Each participant underwent structural MRI scans (T1w MPRAGE and T2-FLAIR), acquired using a 3T MRI scanner located at the Aston Institute of Health *&* Neurodevelopment, Birmingham, UK) (flip angle = 15; dimensions = 176×240×256). From March 2022, the MRI scanner was upgraded from a Siemens TrioTim (TE=(ms) 0.00337; TR(ms)=1.900) to a Siemens MAGNETOM Prisma (TE=(ms) 0.00341; TR(ms)=1.960). Three children with AE and all controls were scanned using the newer Prisma MRI.

Preprocessing of structural MRI (T1w MPRAGE) was done with semi-automated pipelines in FreeSurfer (v6.0^47^): including segmentation of gray matter, white matter and CSF boundaries; brain extraction; quality check for segmentation or surface estimation errors; normalization and automated structural parcellation^48^. Intensity correction was done with non-parametric non-uniform intensity normalization adapted to scans acquired from 3T scanners using the “-3T” flag.^47^ 3D white matter and gray matter surfaces were generated, smoothed and aligned for inter-participant comparability^48^. Pial surface estimation was improved with the contrasts from the FLAIR scans (“-FLAIRpial” argument^47^) when a FLAIR scan was available. Every scan was visually quality-checked for skull-stripping and segmentation errors: manual edits were done in FreeSurfer to remove artefacts, remaining unwanted tissues and to correct surface estimation errors when needed. Scans with excessive noise or artefacts after visual inspection of processed scans were discarded (1 AE and 1 control).

### MEG protocol and processing

#### MEG acquisition parameters

MEG recordings were conducted using the Elekta Neuromag® TRIUX MEG system at the Aston Institute of Health and Neurodevelopment, comprising 306-channels (including 102 magnetometers and 204 planar gradiometers) located in a single-shell magnetically shielded room equipped with MaxShieldTM technology. Recordings were conducted with a 2000 Hz sampling rate, a high-pass filter of 0.1 Hz and a low-pass filter of 330 Hz. Internal active shielding was off. Five head position indicator coils were placed on the participant, three on the forehead and one on each mastoid to record head movements. Co-registration between the MEG and the MRI scans was facilitated using a *Polhemus Fastrak* motion tracker which digitizes the coordinates of each participant’s head shape, starting with three fiducial coordinates (nasion, and bilateral preauricular points), followed by the head position coils and the rest of the head.

#### Resting state protocol

During the MEG recordings, participants underwent a recording at rest, in which they were instructed to sit still in the scanner for a recording period of 6 to 6”30 minutes and to look at a black fixation cross projected on a white background. This was performed in a session in which other, task-based, acquisitions were performed (not reported here).

#### MEG co-registration and source modelling

Each individual’s MRI was co-registered with their MEG recording using BrainStorm (*v.* 3.210818, 18 August 2021^49^), which is documented and freely available for download online under the GNU general public license (http://neuroimage.usc.edu/brainstorm). Co-registration was achieved using fiducial points manually defined on the T1w MRI and refined using the head position indicator coil coordinates. Participant-level anatomy models were imported from the FreeSurfer pipeline described above. The number of vertices used to construct the whole cortical surface generated in FreeSurfer was downsampled to 15000 as per Brainstorm’s recommended default (“good balance between the spatial accuracy of the models and the computation speed”, https://neuroimage.usc.edu/brainstorm/Tutorials/ImportAnatomy).

For cases where this anatomy model was unavailable (for example if an MRI scan contained too many artefacts to generate FreeSurfer meshes or could not be preprocessed), an age-matched paediatric symmetric MRI template^50, 51^ preprocessed with the same FreeSurfer pipeline was used as a substitute. The templates were preprocessed with FreeSurfer to maintain the same parcellation across participants. Template models were used for one AE case and one control participant.

Three-shell realistically-shaped head models were generated for each participant based on their FreeSurfer derived surface-based models, with adaptive integration in *BrainStorm* and the *OpenMEEG* plug-in^52, 53^, using scalp/skull/brain layers with default settings for layer conductivities, vertices and skull thickness. The boundary element method (BEM) was used, as it supports more accurate localization of signal sources compared to sphere-based methods^54^. The source of the signal was reconstructed in each anatomical model using LCMV beamformer, which suppresses external background noise assumed to be captured in a data covariance matrix extracted from each recording, regularized with the median eigenvalues (as recommended^55^).

### Resting state network analysis

Resting state recordings were epoched into non-overlapping trials of 10 seconds using *Fieldtrip* (*v*. 22 January 2021^56^). Signals were filtered with a 4th-order Butterworth zero-phase low-pass filter for better visualization (cut-off: 70 Hz, as in^57^). An independent component analysis (ICA) was run (number of components equal to the number of included channels), to reject heartbeats, eye blinks and eye movement artefacts in all participants. These were visually identified using an in-house script that allows toggling through the components, plotting their topography and signal time-course across all trials. Components containing these artefacts were regressed out of the MEG data. In addition, each individual trial was toggled manually in order to reject trials and channels that contained excessive artefacts caused by muscle activity and SQUID jumps. The cleaned data was then imported into *BrainStorm* (*v.* 3.210818, 18 August 2021^49^) to facilitate connectivity analysis. The number of 10 second epochs that remained after processing were distributed as follows: 37.5±1.6 (IQR = 1) for the Encephalitis group and 37.33±1.1 (IQR = 3) in the Control group.

Before computing connectivity, a weighted average of the trials was obtained and transformed in the time-frequency domain using Hilbert transformation. Frequency bands of interest were delta (1 to 4 Hz) and theta (5 to 8 Hz). Using the Desikan-Killiany atlas, Brainstorm’s scout function was applied to produce the mean time-frequency signal within each parcellated region *before* estimating frequency connectivity (instead of *after* estimating connectivity for each dipole in order to reduce processing time). Connectivity between these regions was computed for each frequency band of interest using Amplitude Envelope Correlation, a measure of temporal evolution of spectral power (envelope), correlated between pairs of orthogonalized signals within separate regions^58^. This method avoids common source contamination and has also been shown to have a superior between-sessions network estimation consistency compared to other measures of connectivity^59, 60^. Two 68×68 connectivity matrices were generated for each epoch and then averaged for each participant; resulting in one average delta connectivity matrix (example given in Supplementary Figure 1) and one average theta connectivity matrix per participant.

The average connectivity matrices were exported individually for each participant in Matlab. Negative correlations were transformed into zeros. The Brain Connectivity Toolbox (BCT *v*. 2019-03-03^61^) was then used to normalize the matrices’ weights into a range from 0 (no connection) to 1 (maximum connectivity). Proportional thresholds were chosen for their higher stability within graph measures^62^; and applied to keep 10% to 30% strongest connections for each frequency matrix (as in previous paediatric MEG research^63^), in intervals of 4%, thus 6 matrices per frequency band. The purpose of looking at the networks with different thresholds is to control for the instability of graph measures across threshold and the arbitrariness of selecting one threshold^64^.

Measures of efficiency and modularity, that reflect how well brain networks integrate and segregate distinct modules to efficiently transmit neuronal information, were chosen. These specific measures were investigated as they were applied in previous AE fMRI research^65, 66^.

For each thresholded matrix, the following measures were computed:

- *Modularity* (*M*), the degree to which the network can be subdivided into non-overlapping groups, maximizing within-group edges, and minimizing between-group edges^61^ (using *Q* maximized modularity).
- *Global efficiency* (*E_glob_*), the average inverse shortest path length between all pairs of nodes.^61^
- Mean *Local efficiency* (*E_loc_*), the global efficiency computed for neighbouring nodes at the level of each node^61^ (with the recommended 2017 BCT function)

### Statistical analyses

#### Multi-threshold permutation correction for network selection

To avoid the issue of multiple comparisons produced by the multiplication of thresholds, as the group difference would need to be assessed for 2 frequency bands * 6 thresholds * 3 graph measures, multi-threshold permutation correction (MTPC) was applied. MTPC was implemented in the *brainGRAPH* package (*v*.3.0.0^67, 68^) in *R* (*v*.4.2.1; R Core Team, 2022). This method allows the identification of a group difference that is stable across thresholds and controls for multiple comparison through permutation correction. The MTPC was successfully applied in other clinical populations with structural diffusion-weighted imaging networks^64, 69–71^, and can be replicated in MEG-derived networks as they are both represented as mathematical graphs. The Desikan-Killiany atlas in brainGRAPH was reordered in order to match the labels of the matrices in Brainstorm. The MTPC computes a test-statistic (here, *t*) on graph measures across groups for each threshold, and permutes group assignments (here, 5000 times). The maximum *t* across all permutations for each threshold are used to establish a critical value at a desired confidence level (here at α = .05). For each threshold, an area-under-the-curve is computed for significant “clusters” where the observed *t* is higher than the critical value. A critical AUC is determined from the mean of the AUCs above critical *t* value of the permuted tests: the output of the MTPC is significant if the AUC of the significant clusters exceeds the critical AUC^64^. In the present study, the MTPC model was one-sided and tested reduced (“less”) connectivity in the AE group.

To account for the multiple MTPC analyses (2 frequency bands * 3 graph measures), the *p* values corrected across all six thresholds in each MTPC (p.fdr values), were corrected again for false discovery rate against the p.fdr values of the other MTPC analyses (6 * 6 p.fdr in total).

For significant differences, a post-hoc exploratory analysis was run to verify a potential difference in overall raw functional connectivity (overall FC), compared across groups using nonparametric permutation tests (t-test with threshold 3.1, two-tailed, 5000 permutations, significant at *p* = .05, component size = extent) in the *Network Based Statistic* toolbox^72^. The point of such post-hoc analysis is to verify whether the network organization differences may simply be explained by the overall difference in FC or, on the contrary, remain different regardless of this overall connectivity contrast. That is because low overall FC can introduce spurious connections within proportional thresholds and in turn influence network metrics^73, 74^. If overall FC differed between groups, a strategy proposed by van den Heuvel *et al.*^74^ was followed if applicable to the data, by establishing overall FC-matched subgroups and rerunning the graph metrics analyses.

A priori power analyses using G*Power^75^, assuming a strong two-tailed individual *t* effect size (*f*^2^ = .35),which is what is required from a relevant biomarker, is observed for the effect of the graph measure, suggest 25 participants are needed to reach a sufficiently high power (1-*β* = .80). Due to recruitment challenges (secondary to the COVID-19 pandemic) that sample size was not reached (N=11, 1-*β* = .41 for the minimum strong effect).

#### Multiple regressions for resting state networks

Regression analyses were computed in *R* (*v*.4.2.1; R Core Team, 2022) using the *car* package^76^. Following the MTPC comparison analyses, graph measures for each participant in the AE group were extracted for the threshold where the group difference was the highest compared to controls, regardless of significance. Graph measures were then separately included in multiple regression analyses predicting Processing Speed Index (PSI) and Working Memory Index (WMI).

Sex was also included in these models. Whilst the WISC-V demonstrates measurement invariance across both males and females,^77^ there is evidence to suggest that cognitive outcomes in this patient population differ as a function of sex. In paediatric ADEM, there is greater risk of neurological poor outcome (including intellectual difficulties) for males.^78^ There are similar trends in paediatric-onset multiple sclerosis, with males being more likely to be cognitively impaired [x],^79^ and experience further decline in cognitive processing speed^80^ and overall cognitive functioning, even at 2-5yrs post-onset.^81^ Given this propensity for sex-differences in cognitive and intellectual outcomes across paediatric neuroinflammatory diseases, sex was therefore included in models as a predictor. The analysis of network associations with cognition without the sex covariate is detailed in the Supplementary material (Supplementary Table 1). Age was not controlled for as the standard WISC scores are already standardized across age-range.

Each linear model was defined as such in R: cognitive measure ∼ graph measure + sex. This amounted to 12 linear models (2 frequency bands * 3 graph measures * 2 cognitive measures). The p-value output for individual graph measure coefficients were corrected for false discovery rate across all the regressions if significant.

A Shapiro-Wilk test was used to assess the normality of the distribution of the cognitive variables. Outliers were defined as three standard-deviations over or under the mean. The assumption of linearity was checked through Residuals vs Fitted scatterplots. The assumption of multicollinearity was tested by computing VIF values (below 10), and the assumption of independence of error residuals was tested with the Durbin-Watson test (non-significant *p* value).

## Results

### Demographics

24 children completed the study, including 12 PAE patients and 12 typically developing children (Table 1). In the PAE patients, the average time from disease onset to scanning was 7.3 years (range 3-15 years). Nine patients were diagnosed with ADEM (four with positive serum MOG-antibodies), and three with autoimmune encephalitis (one with NMDAR-Abs, one with MOG-antibodies and one antibody negative). Three participants had comorbid neurodevelopmental diagnoses; one patient had diagnoses of autism spectrum disorder (ASD), attention deficit hyperactivity disorder, dyspraxia and focal epilepsy; one had epilepsy and one had a diagnosis of ASD. Four patients were on anti-seizure medications. The average Modified Rankin score for the PAE patients was 1.0 (range 0-3).

**Table 1.** Comparison of children with autoimmune encephalitis and typically developing children.

| Child characteristics | Controls<br>N=12 | AE<br>N=12 | Statistics |  |
| --- | --- | --- | --- | --- |
|  |  |  | Test | *p* value |
| Sex (female:male) | 4:8 | 7:5 | $\chi^2 = 1.510$ | .219 |
| Age in years (mean±sd)<br>IQR | 10.5 ± 3.2<br>6.45 | 11.23 ± 3.5<br>6.99 | $t(22) = -$<br>0.503 | .620 |
| Age of onset (mean±sd)<br>IQR | NA | 4.5 ± 3.2<br>3.5 | NA | NA |
| mRS at scan<br>IQR | NA | 0.92 ± 0.9<br>1 | NA | NA |

Note. AE = Autoimmune Encephalitis; mRS = modified Rankin Scale.

### Neuropsychological outcomes

One child with AE did not return for a second visit to complete cognitive assessments. Groups did not significantly differ in Working Memory (*t*(21) = 1.449; *p* = .162; Cohen’s *d* = 0.605), however, the PAE group had a significantly lower average score in Processing Speed (*t*(21) = 2.463; *p* = .023; Cohen’s *d* = 1.028) (Figure 1). Participant scores based on WISC’s age-normalized population categories are depicted in Figure 2. Proportions were compared binary-wise (Extremely low to Low average | Average to Extremely High) between controls and PAE using Fisher’s Exact Test: no significant difference in categories was observed between groups in Processing speed (*p* = .414) or Working Memory (*p* = .193).

**Figure 1.**
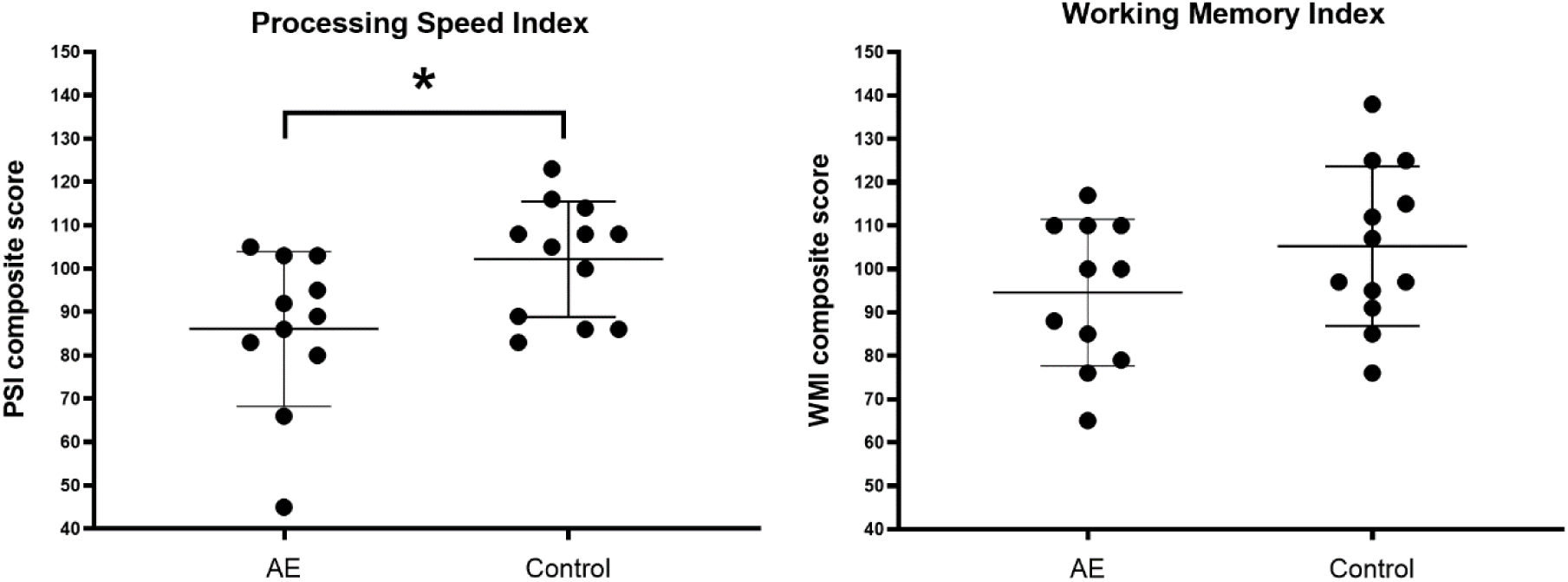
Cognitive outcomes from the neuropsychological assessments across groups. The AE group had a significantly lower average score in Processing Speed (*t*(21) = 2.463; \**p* = .023; Cohen’s *d* = 1.028); measures were obtained with the WISC-V^44^. AE = Auto-immune Encephalitis; PSI = Processing Speed Index; WMI = Working Memory Index;

**Figure 2.**
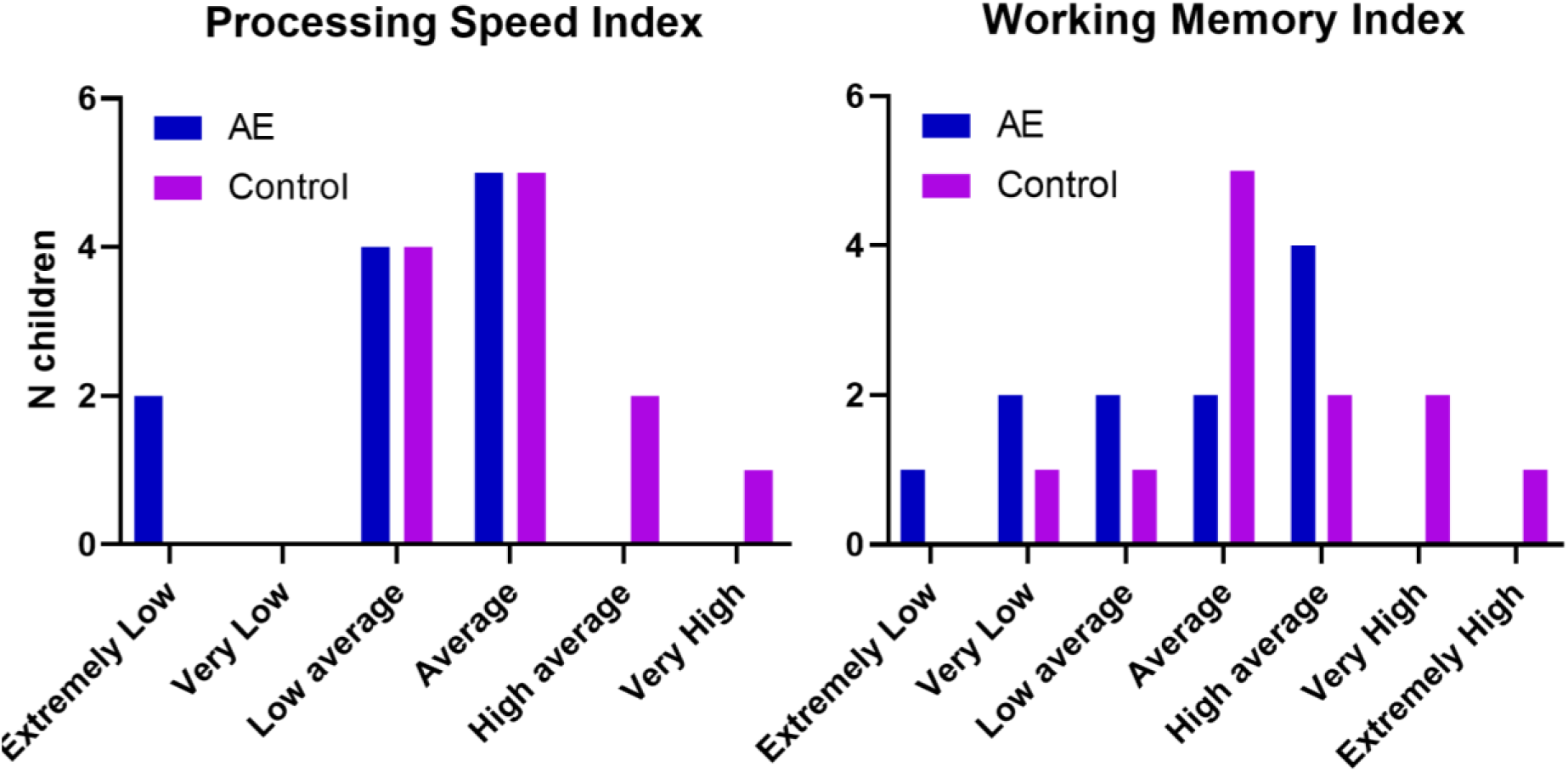
Cognitive scores categorised according to the normative WISC-V classification. AE = Auto-immune Encephalitis. Classification categories and measures were obtained from the WISC-V^37^.

### Resting state network analysis

#### Brain network differences between groups

Resting state MEG recordings were obtained in all 12 controls and 12 children with AE. The MTPC comparison analysis in the delta frequency showed that modularity and global efficiency did not significantly differ, but the mean local efficiency of the AE group was significantly lower than in the Control group (Figure 3). There was no significant difference between the AE group and the Control group in the theta frequency (Figure 4). The group average delta networks are depicted in Figure 5, highlighting regions where efficiency was highest and connections strongest. All significantly different thresholds remained significant at *p* < .05 after all the p values (corrected across thresholds) of the MTPC analyses were corrected again for false discovery rate across all twelve comparison analyses. No difference in overall functional connectivity was observed across groups in either frequency band.

**Figure 3.**
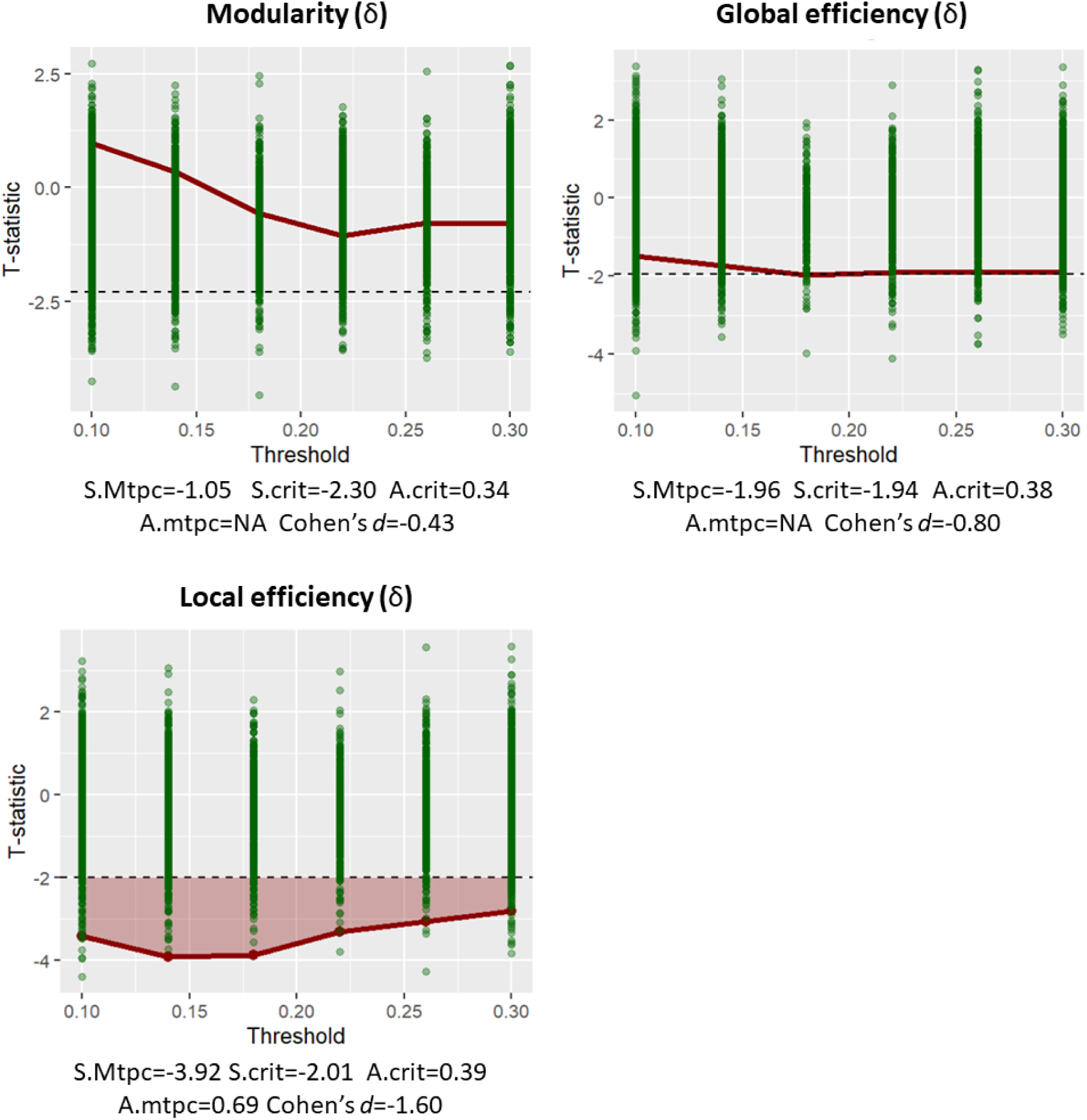
Comparison of delta network measures (1-4Hz) between AE and Control groups. Each graph depicts the network contrast of the AE group in reference to the Control group. The green dots are the maximum null *t* statistics of the 5000 permutations. Red dots are the observed *t* statistics. The dashed line is the critical null statistic (S.crit, top 95th percentile of the null distribution of maximum *t* statistics). The red shaded areas represent clusters of observed statistics above the critical value, whose area-under-the-curve (A.mtpc) is also greater than the mean areas-under-the-curve of the supracritical permuted statistics (A.crit). This means that a lower shaded red bar is a significantly lower network measure compared to the Control group (at *p* < .05). Non-shaded bars are non-significant. The reported effect size *d* is only that of the threshold with the largest difference. S.mtpc = maximum observed statistic; S.crit = critical value of the null max. statistic; A.crit = mean of the supra-critical null AUCs; A.mtpc = AUC value of supracritical cluster.

**Figure 4.**
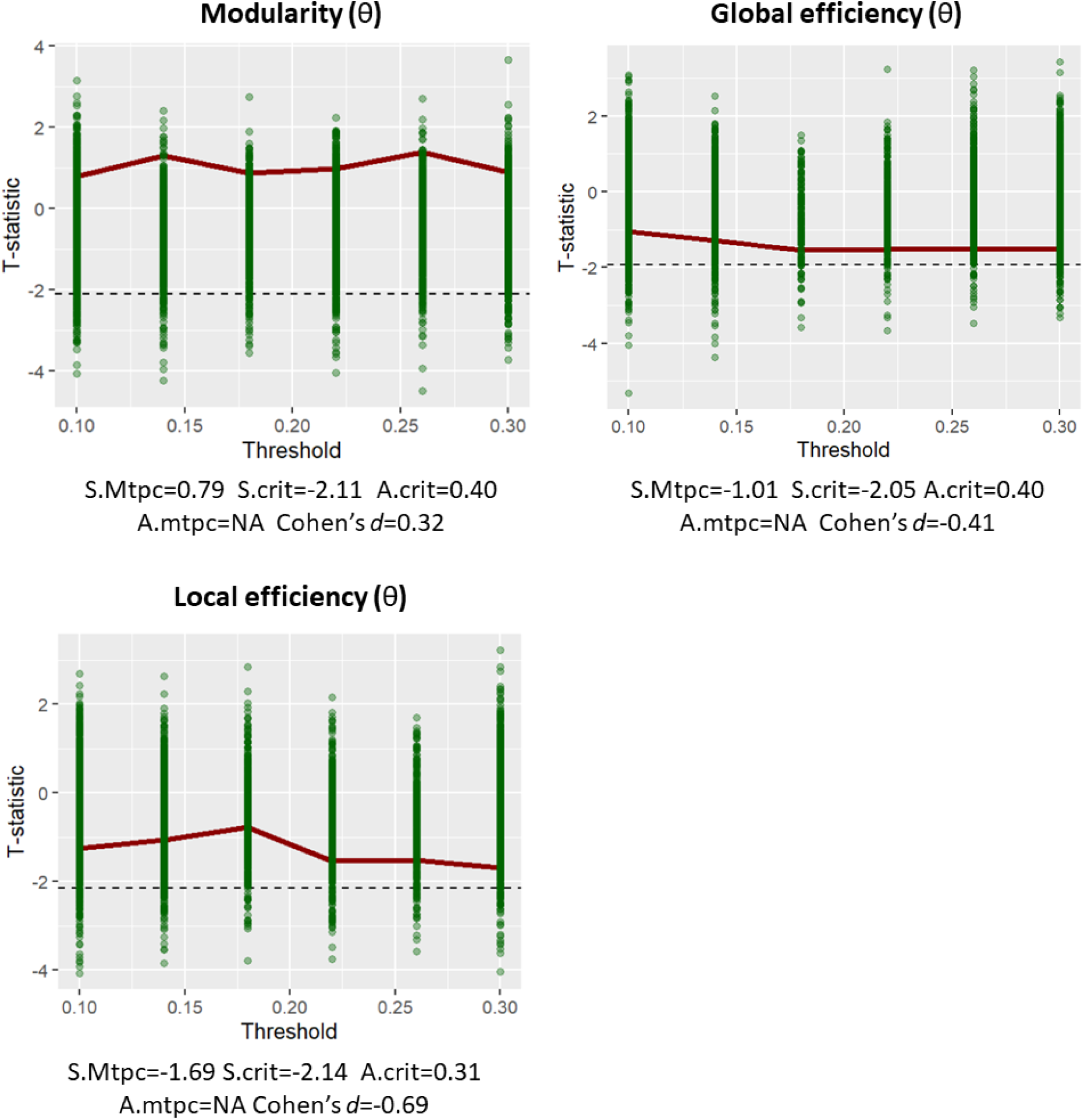
Comparison of theta network measures (5-8 Hz) between AE and Control groups. Each graph depicts the network contrast of the AE group in reference to the Control group. The green dots are the maximum null *t* statistics of the 5000 permutations. Red dots are the observed *t* statistics. The dashed line is the critical null statistic (S.crit, top 95th percentile of the null distribution of maximum *t* statistics). The red shaded areas represent clusters of observed statistics above the critical value, whose area-under-the-curve (A.mtpc) is also greater than the mean areas-under-the-curve of the supracritical permuted statistics (A.crit). This means that a lower shaded red bar is a significantly lower network measure compared to the Control group (at *p* < .05). Non-shaded bars are non-significant. The reported effect size *d* is only that of the threshold with the largest difference. S.mtpc = maximum observed statistic; S.crit = critical value of the null max. statistic; A.crit = mean of the supra-critical null AUCs; A.mtpc = AUC value of supracritical cluster.

**Figure 5.**
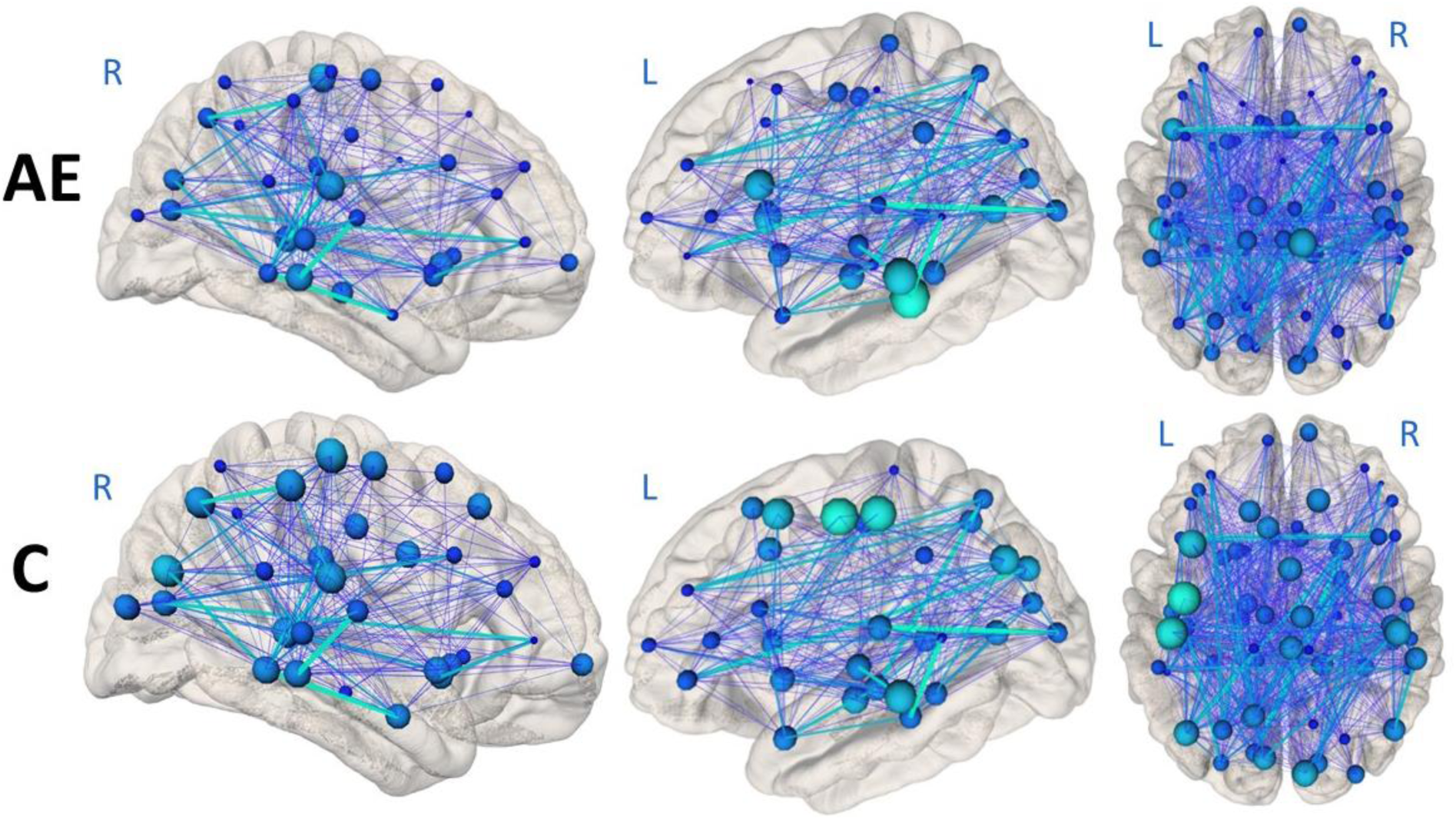
Average delta networks in Autoimmune Encephalitis and Control groups. The figures were produced using the NeuroMArVL web app^69^. 3D surfaces are based on the fsaverage FreeSurfer template. Node size and brightness represent the group average local efficiency per region, and edge thickness and brightness represent the group average strength of the connectivity (not relatively to the other group, on the same scale). The network was thresholded at 14%. AE= Autoimmune encephalitis; C = Controls.

#### Network associations with cognition

Based on the threshold at which group difference had the lowest p-value, the following graph measures were selected as independent variables: theta modularity at threshold 0.1; theta global efficiency at threshold 0.3; theta local efficiency at threshold 0.3, delta modularity at threshold 0.22, delta global efficiency at threshold 0.18, delta local efficiency at threshold 0.14.

One AE case was not included in the regression analyses because the neuropsychological assessments were not completed. This amounted to a group of 11 PAE cases, which is underpowered for a regression analysis with two regressors (1-*β* = .41 for the individual effect coefficient, assuming a large effect size is detected according to a post-hoc estimate in G*Power^75^).

No outliers were detected, and the distribution of the cognitive scores was normal. No multicollinearity between the regressors or significant dependence of error residuals was observed in any model. Their relationship with regressors was linear except for 5 out of the 12 models. Respectively:

- Theta modularity did not have a linear relationship with processing speed index
- Theta global efficiency, theta local efficiency, delta global efficiency and delta local efficiency did not have a linear relationship with working memory index

This violation of linearity could not be solved using log, square root or square transformations of either dependent variables, independent variables or both. The above 5 regressions models were therefore not run. However, given the group difference between AE and controls in delta local efficiency, a non-parametric approach to exploratively test the non-linear relationships was taken using a Spearman correlation (Supplementary Table 2). Delta local efficiency showed a trend towards association with working memory index but this did not reach statistical significance (*p*=0.067).

The overall regression model for theta modularity was not significantly associated with Working memory performance (*F*(2,8)= 3.348; *p* = .088; adjusted *R*^2^ = 0.320). However, theta modularity individually predicted working memory performance but this did not survive false discovery rate correction (*β* = -.781; *t*(8) = −2.588, *p* = .032. All individual effect coefficients are shown in Table 2, model 1). The regression models for theta global efficiency (*F*(2,8)= 0.085; *p* = .919; adjusted *R*^2^ = −0.224) and theta local efficiency (*F*(2,8)= 0.029; *p* = .972; adjusted *R*^2^ = −0.241) did not significantly predict processing speed performance. The regression models for delta modularity did not significantly predict working memory performance (*F*(2,8)= 0.181; *p* = .837; adjusted *R*^2^ = −0.196) or processing speed performance (*F*(2,8)= 0.049; *p* = .952; adjusted *R*^2^ = −0.234). The regression model for delta global efficiency (*F*(2,8)= 0.166; *p* = .850; adjusted *R*^2^ = −0.200) and delta local efficiency did not significantly predict processing speed performance (*F*(2,8)= 0.027; *p* = .974; adjusted *R*^2^ = −0.242).

**Table 2.** Individual effects of delta and theta graph measures on Processing Speed and Working Memory Performance.

| Model | DV | Predictor | *B* | sd<br>error | $\beta$ | *t* | *p* | Confidence<br>interval 95% | |
| --- | --- | --- | --- | --- | --- | --- | --- | --- | --- |
|  |  |  |  |  |  |  |  | Lower | Upper |
| 1 | WMI | $\Theta$ *M* | -493.4 | 190.7 | -0.78 | -2.59 | .032* | -933.1 | -53.7 |
|  |  | Sex | -12.9 | 9.8 | -0.40 | -1.33 | .221 | -35.6 | 9.6 |
| 2 | PSI | $\Theta$ *E<sub>glob</sub>* | -79.7 | 233.5 | -0.15 | -0.34 | .742 | -618.2 | 458.8 |
|  |  | Sex | 5.9 | 15.3 | 0.18 | 0.39 | .705 | -29.2 | 41.2 |
| 3 | PSI | $\Theta$ *E<sub>loc</sub>* | 12.2 | 176.9 | 0.03 | 0.07 | .947 | -395.9 | 420.3 |
|  |  | Sex | 2.3 | 13.8 | 0.07 | 0.17 | .871 | -29.4 | 34.05 |
| 4 | WMI | $\delta$ *M* | -164.7 | 273.9 | -0.23 | -0.60 | .564 | -791.4 | 476.1 |
|  |  | Sex | -3.1 | 12.2 | -0.10 | -0.25 | .805 | -31.2 | 25.0 |
| 5 | PSI | $\delta$ *M* | -62.8 | 293.4 | -0.08 | -0.21 | .836 | -739.3 | 613.7 |
|  |  | Sex | 1.7 | 13.0 | 0.05 | 0.13 | .900 | -28.4 | 31.7 |
| 6 | PSI | $\delta$ E.Glob | 99.9 | 190.0 | 0.19 | 0.53 | .613 | -338.3 | 539.2 |
|  |  | Sex | 0.6 | 12.6 | 0.02 | 0.05 | .963 | -28.4 | 29.5 |
| 7 | PSI | $\delta$ E.Loc | -5.172 | 171.6 | -0.014 | -0.030 | .977 | -400.0 | 390.6 |
|  |  | Sex | 3.057 | 15.43 | 0.089 | 0.198 | .848 | -32.53 | 38.64 |

Note. DV = Dependant variable. WMI = Working Memory Index. PSI = Processing Speed Index. M = Modularity. *E<sub>glob</sub>* = Global efficiency. *E<sub>loc</sub>* = Local efficiency. Cognitive measures were the composite scores from the WISC-V; \* $P < .05$ (fdr corrected $p = 0.224$ )

## Discussion

This study employed advanced network analyses of MEG recordings to investigate whether disruption to functional connectivity was associated with cognitive outcome in paediatric autoimmune encephalitis. MEG resting-state connectivity in delta and theta frequency bands were analysed in a cohort of children with AE and a group of similarly aged typically developing controls. Metrics describing the organisation of the functional connectivity data were included in linear (and non-linear) models to predict processing speed and working memory as these are domains known to be affected in PAE. Local efficiency within delta networks was lower on average in AE compared to controls (and trended towards an association with working memory index). Modularity in theta networks significantly predicted lower working memory performance in AE. This is the first study to demonstrate feasibility, validity, and potential *predictive* validity of MEG-based functional connectivity analyses in a cohort of children with AE.

In this current study, a proportion of PAE patients had an identifiable cognitive impairment in processing speed (33%) and working memory (16.7%), consistent with other studies^5, 82–84^. It is important to note here that the PAE patient cohort were considered to have a “good” outcome on average (mRS score 0 to 2^4^) based on the Rankin scale’s measure of disability, with only one child having moderate disability with a mRS of 3. Processing speed was significantly lower on average in the PAE cohort compared to the controls and is one of the most common difficulties observed in paediatric NMDARE and ADEM^5, 82, 85^. This suggests PAE patients may be more likely to struggle with responding rapidly to task-relevant information, even some years after disease onset. The use of resting state neuroimaging paradigms in this population is therefore highly relevant, being easier to undertake by these children who may struggle with the demands of an interactive task or paradigm whilst in the MEG.

Resting-state network connectivity and topology were assessed within delta and theta frequency bands because of their relevance for cognition^41, 42^ and their previously observed alterations in adult AE studies (with the exception of Miao et al. reporting a MEG study on a 16 year old patient)^27, 37, 57, 86^.

In the delta frequency, neither modularity nor global efficiency differed from controls, but local efficiency was significantly lower compared to controls. This suggests that while a globally efficient topology and modularity of the overall functional brain network is preserved, the local topology is altered. When considering brain regions that form local neighbourhoods and are functionally ‘connected’ (these are not necessarily local in the sense of spatial location in the brain), information transmission between these regions is less efficient than would typically be expected in the healthy brain. The delta band specificity may be linked to the abnormalities that are commonly reported in AE patient EEG. For example, extreme delta brush (delta activity with superimposed bursts of beta frequency), which can also be observed with MEG^27, 3^ and generalised rhythmic delta activity are common observations in NMDARE^39^. Slowing of brain activity (in the delta frequency range) is frequently noted in ADEM^87, 88^ and NMDARE^89^ patient EEG. This present study suggests slow-wave delta activity may be affected in PAE years after onset and could represent a good candidate biomarker for future research.

In the theta frequency, modularity, global efficiency and local efficiency were not significantly different in PAE compared to controls. These measures estimate the extent to which the brain is efficient in transmitting information across the whole network while maintaining efficient and distinct local groups of brain activities.

With regards to correlation with cognitive outcome, one graph measure – theta modularity-returned a significant result (but did not survive correction for multiple comparisons). Theta modularity was significantly associated with lower working memory performance, i.e., increased modularity in the theta band was associated with lower working memory performance. While the failure to survive correction may be explained by lack of statistical power, such a result, if replicated in larger studies, suggests that the more theta activity tends to form individual and distinct modules within the network, the harder it is for children with AE to keep information in mind and manipulate it. This may reflect a narrowing of a normally widely distributed cognitive network underlying working memory functioning. Recent research has shown that theta connectivity in specific edges were correlated negatively to working memory in paediatric temporal lobe epilepsy^90^ supporting our observation that alterations in typical patterns of theta activity may interfere with working memory function in children with neurological disease. Theta modularity may represent a sensitive measure to detect differences in working memory in PAE^5, 82^.

None of the other graph measures significantly predicted processing speed or working memory when a valid statistical model was used. Adult AE studies have found associations between fMRI connectivity in regionally defined networks with working memory and processing speed^18, 91^. Previous fMRI studies in NMDARE observed a higher connectivity to neighbouring nodes in the right lateral orbital gyrus compared to controls^66^, and lower clustering coefficient and local efficiency in the left insula^65^, which suggests differing local areas within a network could be identified in future studies. The present study lacked statistical power to investigate specific regions of interest and was focused on global and average measures of network efficiency.

## Limitations

A small sample size (a result of the COVID pandemic, condition rarity and single-centre recruitment), is the main study limitation. The present findings must therefore be interpreted with caution and cannot be extrapolated to the entire population of children with AE. However, they do offer compelling preliminary findings and provide direction for future larger-scale studies. Additionally, this “real-world” cohort was heterogeneous in terms of subtypes of AE and was too small to be subdivided.

In terms of analysis, adult atlases were used for parcellation of the MRI acquired brain surfaces, because no atlas specific to the age-range of the cohort was available in FreeSurfer. The selected atlases were not standardized for children of that age range and therefore may lack accuracy. However these atlases have been validated in children from 4 to 11 years^92^ and are routinely used in paediatric samples.^93^ There were also manual processes, such as manual edits to correct for MRI artefacts, or manual fiducial positioning of the MEG coordinates along the MRI scans, which could not be automated with the *Neuromag* coordinate system. Manual intervention may have introduced a level of bias and to mitigate this, the guidance detailed in the FreeSurfer online resources was followed (https://surfer.nmr.mgh.harvard.edu/fswiki/FsTutorial/TroubleshootingData). Although maxfiltering was not used on MEG resting state recordings, and no principal component analysis was used, an independent component analysis was deemed necessary to obtain a sufficiently clean data for further analysis. This involves the risk of the phase being distorted as documented in some EEG simulations^94, 95^. Because spurious changes in phase may occur, so do spurious indices of phase coherence when measuring connectivity^94^. This was further demonstrated by phase distortion observed in segments that were specifically artefact-free after an ICA run on overall EEG recordings^95^. Because the connectivity analysis of the present study relied on amplitude envelope correlation rather than phase-locking connectivity, it may be less affected by phase distortion (given the envelope is a broader measure of the signal). The extent to which such distortion impacts MEG data is unknown, and if it exists, there is a risk that artificial correlations between signals appeared as a result of random distortions. Thresholding may have however minimized the likeliness of including spurious connections. Signal decomposition in paediatric MEG data still appears to be the most efficient way to address artefacts, given manual inspection and attempts to minimize motion during the recordings were not sufficient to produce clean data in the present paediatric cohorts.

## Conclusion

This study reinforces previous findings that children with AE have ongoing residual cognitive difficulties in the long term, with lower performance in processing speed compared to typically developing children, despite being classified as having a “good” medical outcome. Resting-state MEG recordings indicated lower local efficiency within delta frequency networks of children with AE and that higher levels of modularity within the theta-frequency resting-state network may be associated with lower working memory. Future studies will benefit from larger sample size and newer neuroimaging approaches, for example using OPM-MEG, guided by the measures investigated in the present study. The preliminary data presented supports MEG as an appropriate and feasible technique to characterise functional dysconnectivity in paediatric autoimmune encephalitis.

## Data Availability

The data that support the findings of this study are available from the corresponding author, upon reasonable request.

## Acknowledgements

Acknowledgement is given to Professor Stefano Seri, Drs Oana Badaru, Jan Novak, Adam Shephard, Katherine Ellis, and Miss’ Rochelle Hamilton and Rebecca Collings for their involvement in broader aspects of the current program of work in which this project falls. Special thanks to Ms. Lydiah Makusha for valuable help with recruitment into the study. We would also like to thank the West Midlands Clinical Research Network for their study support and Encephalitis International for supporting CB and the research team.

## Supplementary material

### Methods

**Supplementary Figure 1.**
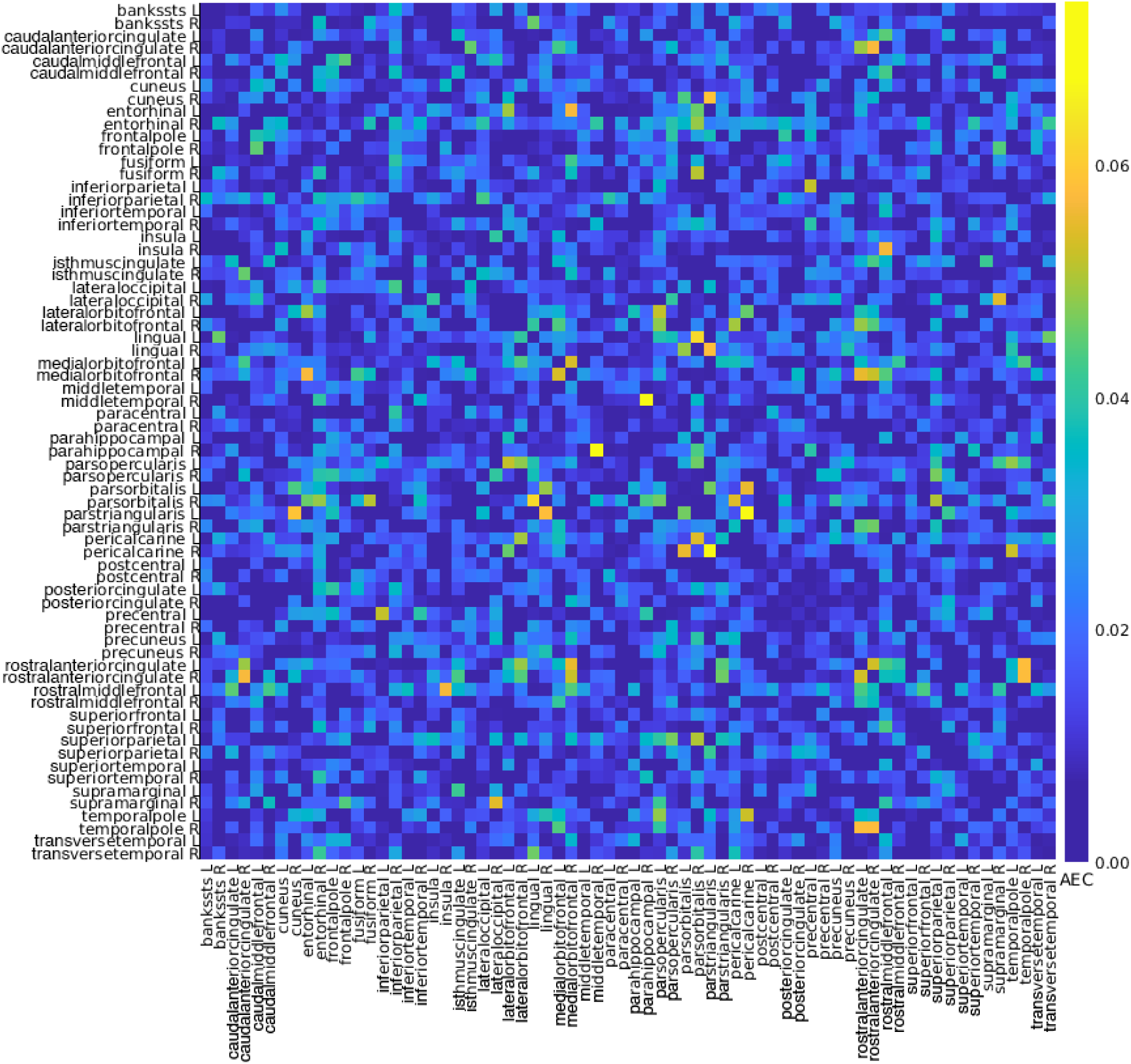
Delta connectivity matrix estimated in a participant. AEC = Amplitude Envelope Correlation. The figure was produced with Brainstorm^44^. The matrix depicts the level of connectivity between each of the 68 brain regions parcellated with the Desikan-Killiany atlas.

### Results

#### Network associations with cognition without the sex covariate

To investigate the effect of the graph metrics on the WISC measures on their own, separate models were also tested excluding sex as a control variable (Supplementary Table 1). The regression model for theta modularity was not significantly associated with Working memory performance (*F*(1,9)= 4.552; *p* = .062; adjusted *R*^2^ = 0.262). The regression models for theta global efficiency (*F*(1,9)= 0.017; *p* = .898; adjusted *R*^2^ = −0.109) and theta local efficiency (*F*(1,9)= 0.033; *p* = .861; adjusted *R*^2^ = −0.107) did not significantly predict processing speed performance. The regression models for delta modularity did not significantly predict working memory performance (*F*(1,9)= 0.332; *p* = . 579; adjusted *R*^2^ = −0.071) or processing speed performance (*F*(1,9)= 0.092; *p* = .769; adjusted *R*^2^ = −0.100). The regression model for delta global efficiency (*F*(1,9)= 0.370; *p* = .558; adjusted *R*^2^ = −0.067) and delta local efficiency did not significantly predict processing speed performance (*F*(1,9)= 0.016; *p* = .901; adjusted *R*^2^ = −0.109).

**Supplementary Table 1.** Individual effects of delta and theta graph measures on Processing Speed and Working Memory Performance without sex as a control variable.

| Model | DV | Predictor | *B* | sd error | $\beta$ | *t* | *p* | Confidence interval 95% | |
| --- | --- | --- | --- | --- | --- | --- | --- | --- | --- |
|  |  |  |  |  |  |  |  | Lower | Upper |
| 1 | WMI | $\Theta M$ | -366.36 | 171.71 | -0.579 | -2.134 | .062 | -754.80 | 22.08 |
| 2 | PSI | $\Theta E_{glob}$ | -22.82 | 174.43 | -0.043 | -0.131 | .898 | -417.40 | 371.75 |
| 3 | PSI | $\Theta E_{loc}$ | 26.55 | 146.40 | 0.060 | 0.181 | .861 | -304.62 | 357.72 |
| 4 | WMI | $\delta M$ | -137.85 | 239.22 | -0.189 | -0.576 | .579 | -678.99 | 403.29 |
| 5 | PSI | $\delta M$ | -77.48 | 255.42 | -0.101 | -0.303 | .769 | -655.28 | 500.33 |
| 6 | PSI | $\delta E_{Glob}$ | 102.91 | 169.20 | 0.199 | 0.608 | .558 | -279.86 | 485.67 |
| 7 | PSI | $\delta E_{Loc}$ | 16.06 | 126.68 | 0.042 | 0.127 | .902 | -270.52 | 302.63 |

PSI processing speed index WMI working memory index

**Supplementary Table 2.** Exploratory analysis of relationships between predictor variables and dependent variables that could not be tested in the regression, using Spearman correlation (linear relationship not assumed).

| Relationship | S | *p*-value | *rho* |
| --- | --- | --- | --- |
| Theta modularity + PSI | 263.1 | 0.563 | -0.196 |
| Theta global efficiency + WMI | 234.1 | 0.851 | -0.064 |
| Theta local efficiency + WMI | 195.7 | 0.747 | 0.110 |
| Delta global efficiency + WMI | 218 | 0.979 | 0.009 |
| Delta local efficiency + WMI | 94.6 | 0.067 | 0.570 |

PSI processing speed index WMI working memory index

